# Trends in anti-hyperglycaemic drug usage among Danish type 2 diabetes patients on second-line treatment and beyond: A nationwide cohort study

**DOI:** 10.1101/2025.06.20.25328883

**Authors:** Henrik Vitus Bering Laursen, Flemming Witt Udsen, Morten Hasselstrøm Jensen, Peter Vestergaard, Søren Paaske Johnsen

**Affiliations:** Danish Center for Health Services Research, Department of Clinical Medicine, Aalborg University, Aalborg, Denmark; Steno Diabetes Center North Denmark, Aalborg, Denmark; Department of Endocrinology, Aalborg University Hospital, Aalborg, Denmark; Department of Clinical Medicine, Aalborg University Hospital, Aalborg, Denmark; Department of Health Science and Technology, Aalborg University, Aalborg, Denmark; Data Science, Novo Nordisk A/S, Søborg, Denmark

## Abstract

2

**OBJECTIVE:** The paradigm shift introduced by the glucagon-like peptide-1 (GLP1) receptor agonist and sodium-glucose co-transporter-2 (SGLT2) inhibitor classes of drugs has led to changes in treatment practices in Denmark. The aim of this study was to investigate what these changes were before and after the implementation of the 2018 guidelines in Denmark.

**RESEARCH DESIGN AND METHODS:** Registry data on prescriptions was linked with other registries to create a descriptive overview of treatment patterns was carried out for a nationwide cohort of all individuals receiving at least two non-insulin anti-diabetic drugs (NIADs) between 2012 and 2021.

**RESULTS:** A cohort of 66,188 individuals was identified. The treatment patterns reflected significantly higher use of SGLT2 and GLP1 over time, eclipsing all other drugs, except for insulin, at later treatment stages. First-line use of SGLT2 and GLP1 also became higher over time, as did their likelihood of prescription at later stages. Their combined use rivaled insulin usage at similar stages by 2021. First-line use for women were high in the GLP1 group.

**CONCLUSIONS:** Our study finds that GLP1 and SGLT2 are used extensively for individuals who have tried multiple NIADs and may delay the use of insulin. Furthermore, there is an increased trend in their first-line use.

**Key messages:**

- What is already known on this subject

○ Increase in use of SGLT2 and GLP1 is documented, but changes in treatment patterns specifically for a cohort of people with type 2 diabetes before and after the publication of the 2018 treatment guidelines has not.
- What did we find?

○ A marked increase in SGLT2 and GLP1 utilization, an indication that insulin initiation is postponed following this, and a preference for GLP1 in younger people in the cohort.
- What are the implications of our findings?

○ Introduction of SGLT2 and GLP1 has led to major changes in the treatment practice of type 2 diabetes. Systematic population-based monitoring is highly warranted to ensure that treatment practice remain compliant with clinical guidelines recommendations.

---

The 2018 ADA/EASD consensus report (1) and its subsequent updates (2) place glucagon-like peptide-1 (GLP1) receptor agonists and sodium-glucose co-transporter-2 (SGLT2) inhibitors at the center of their recommendations for the treatment of type 2 diabetes due to their anti-hyperglycaemic and cardiorenal protective effects. These two classes, along with dipeptidyl peptidase-4 (DPP4) inhibitors, are commonly referred to as newer non-insulin anti-diabetic drugs (nNIADs). Danish guidelines have been aligned with EASD/ADA guidelines (3,4), and the use of nNIADs has increased rapidly in Denmark (5,6), as well as internationally (7–24).

The ADA guidelines published in 2022 (25) recommend using GLP1 and SGLT2 for patients with cardiorenal risk factors, independent of glycemic management, which promotes these drugs to the de facto first-line option for these patients. This challenges the longstanding consensus on using metformin as the first-line treatment. Discussions regarding the balance between the high benefits and costs of these drugs have been ongoing in the US, with calls for price negotiations, especially for GLP1 (26).

The use of SGLT2 and especially GLP1 has been called excessive and outside the restrictions of reimbursement schemes (27,28), although others have pointed out that off-label use of GLP1 is minor (29). A proper discussion on the consequences of their increased use relies on real-world data to monitor changes in treatment. A study on the increased utilization, resulting treatment patterns, and characteristics of individuals undergoing these treatments has not yet been conducted in a Danish context.

This study aimed to use Danish registry data to create an overview of the changes in treatment patterns since 2012. Temporal changes in treatment patterns for NIADs and insulin are investigated for the target demographic as described in the 2018 guidelines for the treatment of type 2 diabetes.

## 3 Research Design and Methods

### 3.1 Study Design

To investigate the changes in treatment patterns over time, we established a registry-based cohort of all Danish individuals with type 2 diabetes who received at least second-line NIAD treatment between 2012 and 2021. Treatment patterns were examined through changes in drug utilization, treatment stages, and the most frequent treatment stage pathways. Baseline characteristics were provided for adding context to the demographics whose treatment patterns were explored. This study adhered to the RECORD (Reporting of studies Conducted using Observational Routinely collected health Data) guidelines (30).

### 3.2 Setting and Data Sources

Danish registry data has long been utilized to produce high quality research. The registries are all linked through a personal identification number given at birth or upon immigration (31,32). This linkage facilitates nationwide cohort studies with virtually complete long-term follow-up, ensuring high accuracy and validity of information (31,32). The primary data source used to establish the cohort was the Danish National Prescription Database (DNPD) (33). This database includes nationwide information on redeemed prescriptions dispensed by Danish community pharmacies since 1996. Anatomical Therapeutic Chemical (ATC) codes on redeemed prescriptions were used as proxies for treatment with the respective drugs, and prescription dates served as inclusion times. Drug utilization was expressed as defined-daily dosages (DDD). Information on sex and age was retrieved from the Danish Civil Registration System (31). Data access can be granted with affiliation with a Danish research organization, through Statistics Denmark.

### 3.3 Study population

Individuals were eligible for inclusion in the cohort if they had received second-line treatment for type 2 diabetes at any point during the study period. This was defined as having prescriptions redeemed for at least two different NIADs, and least two prescriptions of any NIAD different from the first. The inclusion period for both cohorts started in 2012 as this was the first year where all nNIAD classes were used in Denmark and ended in 2021 as this was the end of data availability. An early inclusion period from 2012 to 2018 and a late inclusion period from 2019 to 2021 were defined. To avoid including individuals with type 1, those with any prescription of insulin before baseline. Additionally, those with any NIAD prescription before 2012 were excluded. The cohort was followed from the date of first prescription of any NIAD.

### 3.4 Variables

Baseline characteristics for the cohort included age, sex, inclusion period, and the first prescribed drug class. The latter was defined as the first redeemed prescription of a non-insulin antidiabetic drug (NIAD) the patient received. The purpose was to examine changes in general characteristics and the drug classes used as first-line therapy. These characteristics were compared before and after 2019, since the 2018 guidelines were published half-way through 2018 in Denmark (3). Temporal treatment pattern trends were investigated through Defined Daily Doses (DDD) and treatment stages. DDD, as defined by the World Health Organization (WHO), represents the expected average maintenance dose when the drug is used for its main indication in an adult population. This measure was used to illustrate changes in drug utilization.

To approximate changes in treatment stage patterns, ranging from first-line to sixth-line choices, the prescribed drugs were ordered according to their sequence of introduction for each individual patient. The frequency with which each drug appeared as the first to the sixth was then calculated. This counting was performed at the product level (ATC level 5). For instance, if the first prescription was for metformin and the next distinct prescription was for a sulfonylurea, metformin was classified as first-line, and sulfonylurea as second-line. These instances were then aggregated at the class level. Consequently, if the second unique drug was within the same class as the third, it contributed to the counts of that drug class as both second-line and third-line. No distinction was made between whether the introduction of a new class was due to a switch or an addition. All insulin classes were merged into one.

The most frequent treatment stage pathways for all patients were based on the treatment stages above. An example pathway would be “metformin → GLP1 → Insulin”. This represents a patient who received metformin as the first drug, after which the second unique drug was GLP1, and insulin the third, over the entire follow-up period.

### 3.5 Statistical methods

For the baseline characteristics, continuous variables were presented with mean and standard deviation (SD), and categorical variables were presented with counts and proportions. The proportion of total DDDs for each class was presented as percent in bar graphs. DDDs per 1000 inhabitants per day was based the entire population of Denmark and presented in line graphs. This was calculated for every year with the following equation:

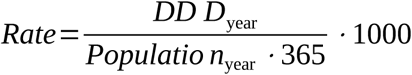

The changes in treatment stages over time, and treatment stage pathways in the early and late periods, were summarized with counts and proportions and presented in bar charts and a table, respectively. All analyses were performed using R (version 4.3.2).

## 4 Results

### 4.1 Study population

From the DNPD we identified 212.752 individuals receiving NIADs since 2012, out of which 66.188 fitted the inclusion criteria for the cohort. See Figure 1.

**Figure 1:**
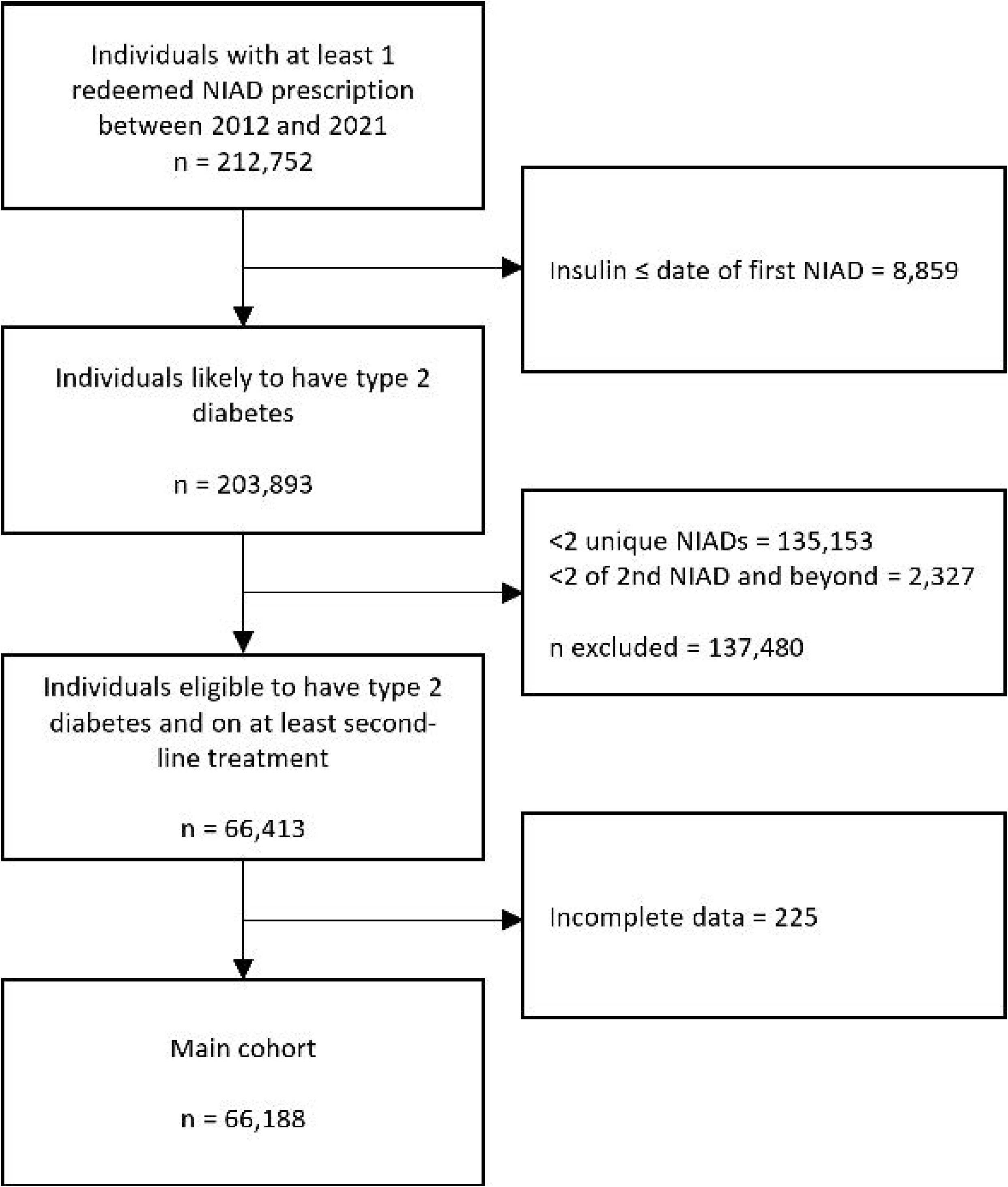
Flow diagram showing the selection of the cohort. NIAD = non-insulin anti-diabetic drug, nNIADs = newer NIADs.

### 4.2 Cohort characteristics

The baseline characteristics of the cohort are detailed in *Table 1*. The majority (75.2%) of the cohort was aged between 45 and 74 years. In both the early and late periods, males constituted the majority (58.8% and 60.6%, respectively). Metformin as the first-line drug showed a decline over the two periods, from 94.6% to 87.6% in women and from 95.6% to 92.1% in men. Conversely, GLP1 as first-line drug increased by 6.6 percentage points in women and 2.7 in men, marking the second-largest shift observed.

**Table 1:**
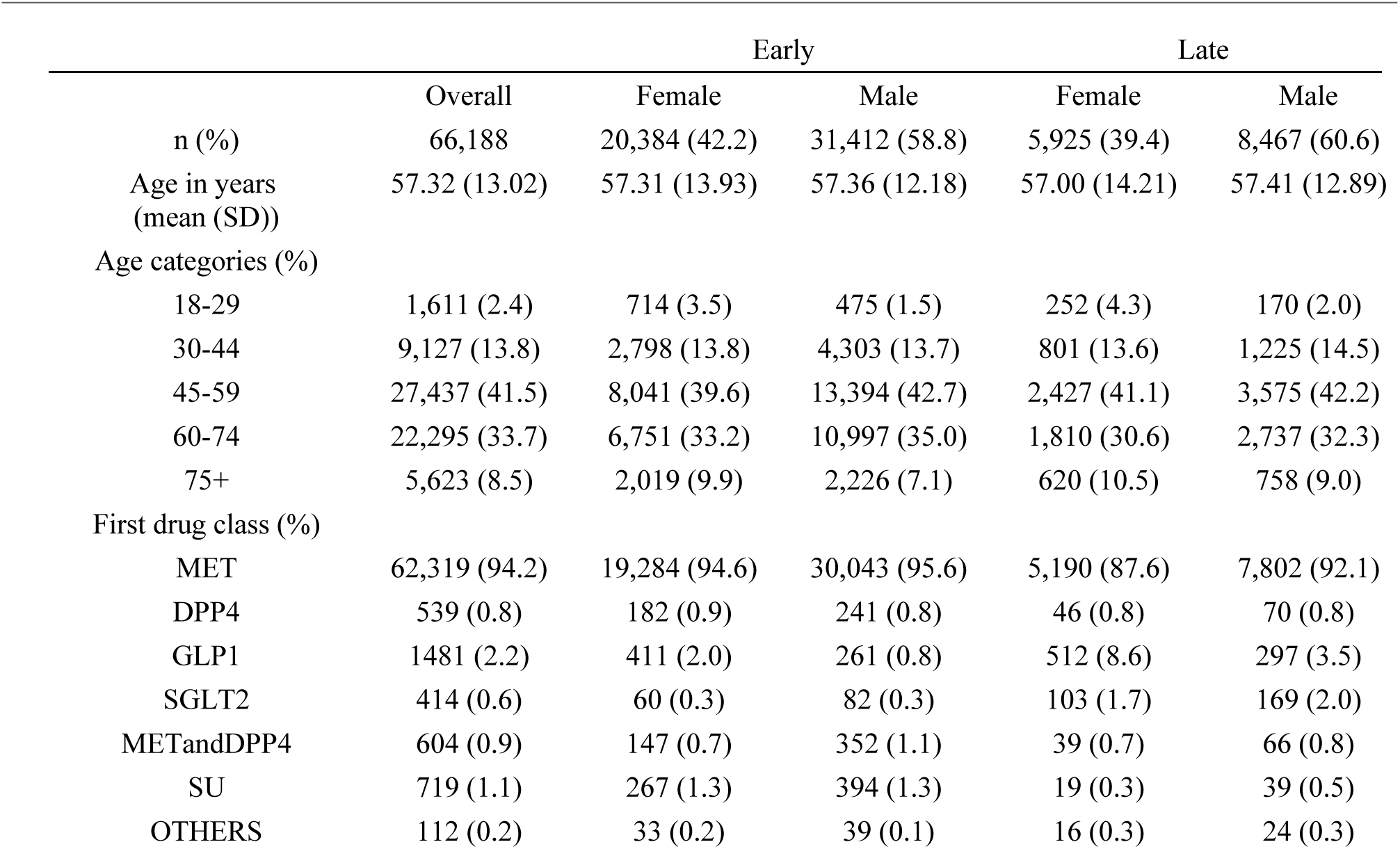
Baseline characteristics of the cohort. Abbreviations: Early/Late = period of inclusion, based on first drug prescribed, for cohort before and after 2019, respectively. MET = metformin, DPP4 = dipeptidyl peptidase-4 inhibitors, GLP1 = glucagon-like peptide-1 receptor agonists, SGLT2 = sodium glucose co-transporter-2 inhibitor, METandDPP4 = metformin and DPP4 combination, SU = sulfonylurea, OTHERS = all other non-insulin anti-diabetic drugs.

### 4.3 Drug utilization

The utilization rate and proportion of total DDD in percent of all NIAD classes and insulin over time are presented in Figure 2. Since 2012, metformin utilization has slowly decreased in proportion, with a notable decline in rate occurring for the first time between 2020 and 2021. For DPP4, the proportion increased until 2016 and then declined, whereas SU began decreasing in 2014. The rates of both DPP4 and SU increased slowly from 2012, and remained constant for DPP4 from 2020, and for SU from 2017. In contrast, GLP1 receptor agonists and SGLT2 have steadily increased in proportion since 2013, with SGLT2 surpassing in proportion since 2017. Rates for SGLT2 and GLP1 increased markedly since 2017 and 2019, respectively. Insulin usage as a proportion increased slightly until 2017, after which it remained constant, while the rate has been slowly increasing.

**Figure 2:**
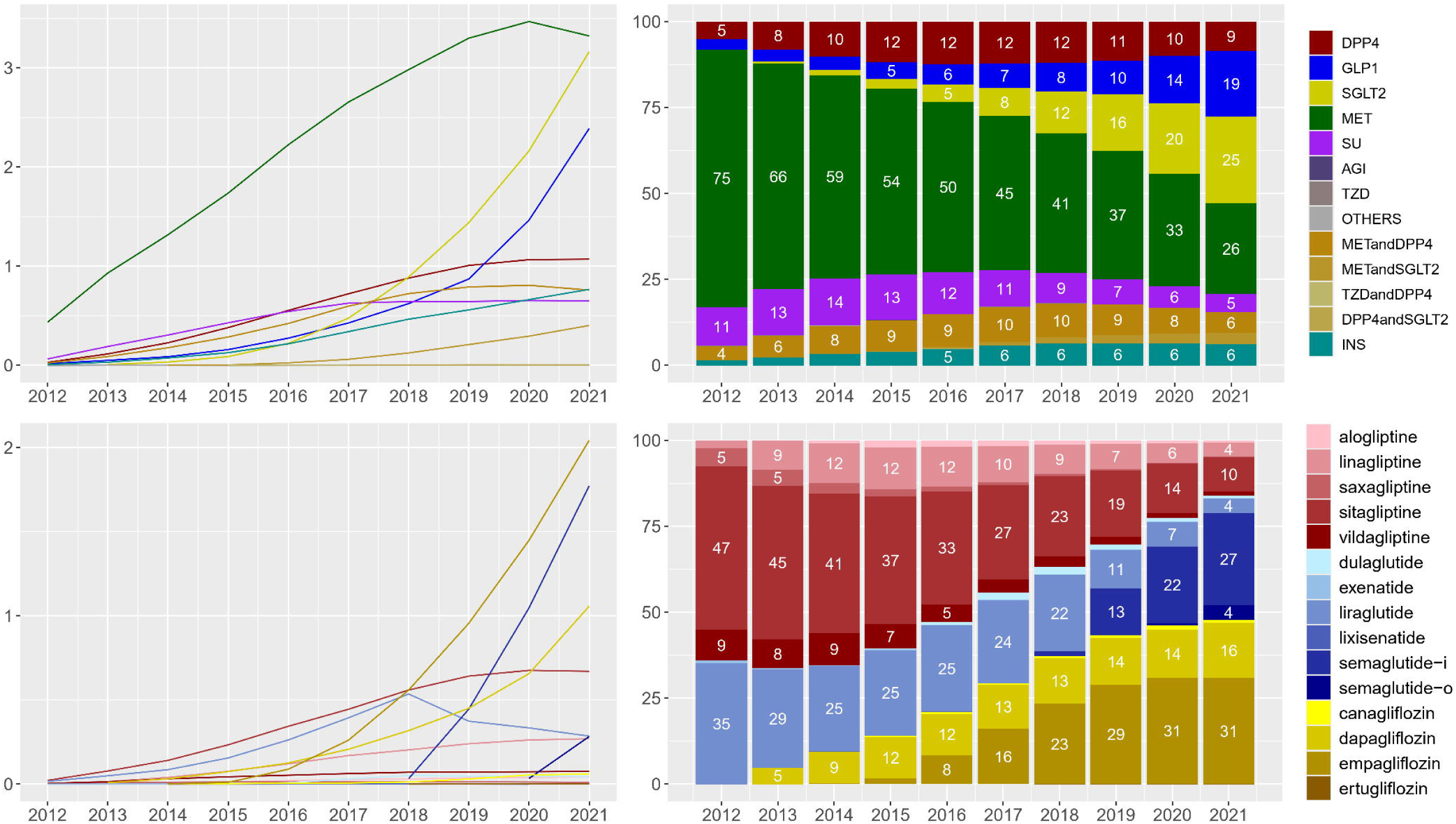
Line charts (DDDs per 1000 inhabitants per day) and stacked bar chart (proportion of total DDD) for the GLP1, SGLT2 and DPP4 drug classes (top row) and products within those classes (bottom row), respectively. MET = metformin, SU = sulfonylurea, DPP4 = dipeptidyl peptidase-4 inhibitors, GLP1 = glucagon-like peptide-1 receptor agonists, SGLT2 = sodium glucose co-transporter-2 inhibitor, TZD = thiazolidinedions, AGI = alpha-glucosidase inhibitor, INS = insulin, METandDPP4 = metformin and DPP4 combination, OTHERS = all other anti-hyperglycaemic drugs.

### 4.4 Treatment Stages

The proportions of drug classes used in treatment stages changed over time, as shown from the first- to the sixth-line treatment stage in Figure 3. In 2012, the first drug introduced was almost exclusively metformin, at 96.5%, with only 0.5% receiving GLP1 and none receiving SGLT2 as first-line treatments. By 2021, the first-line use of metformin decreased to 86.1%, while the first-line use of GLP1 and SGLT2 increased to 3.7% and 8%, respectively.

**Figure 3:**
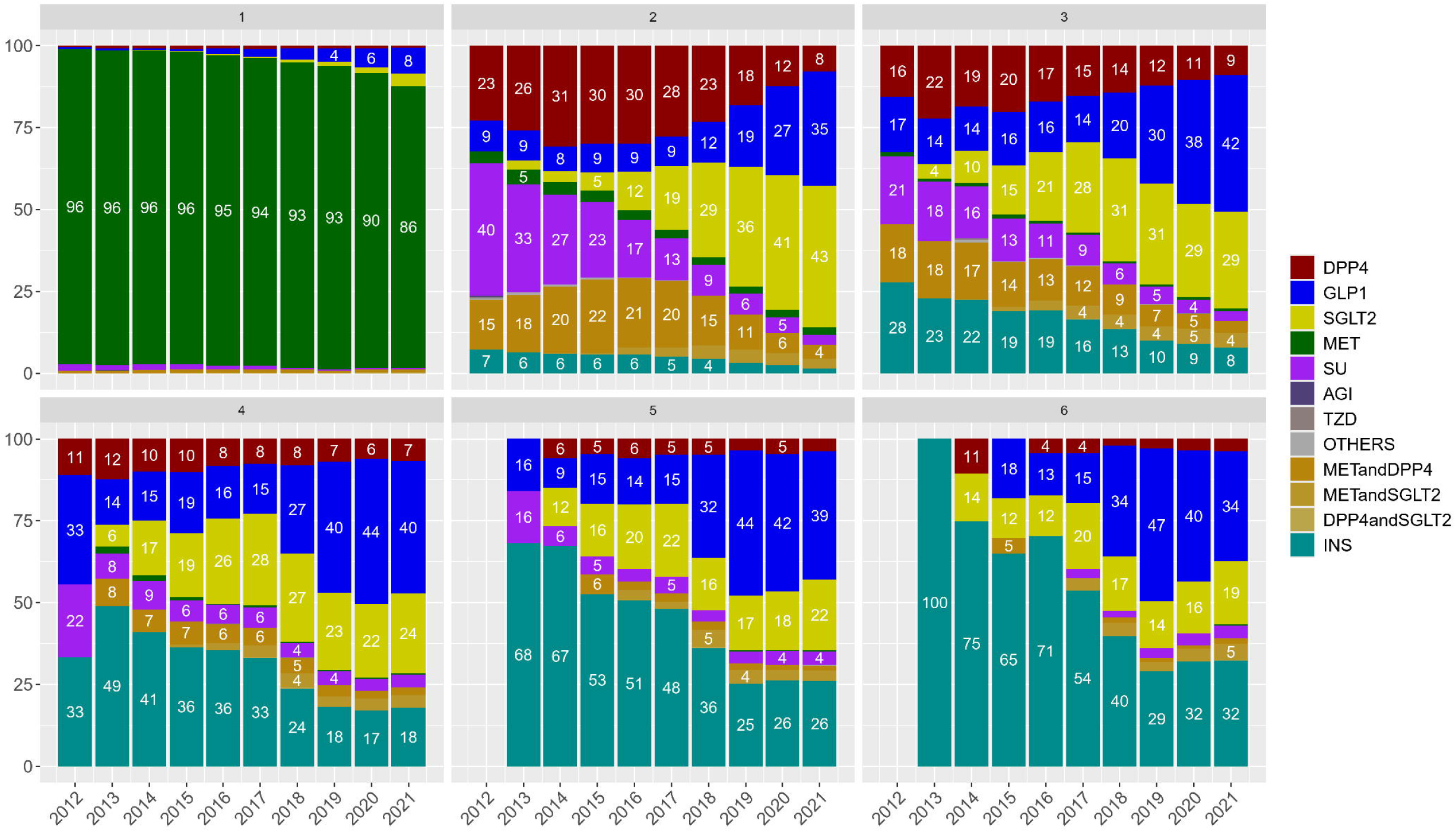
Depicted here are the proportions of each drug class at each treatment stage for the cohort, i.e. the first to the sixth introduced drug for each individual patient, over time. Omitted bars in 2012 are due to insufficient or lacking data. MET = metformin, SU = sulfonylurea, DPP4 = dipeptidyl peptidase-4 inhibitors, GLP1 = glucagon-like peptide-1 receptor agonists, SGLT2 = sodium glucose co-transporter-2 inhibitor, TZD = thiazolidinedions, AGI = alpha-glucosidase inhibitor, INS = insulin, METandDPP4 = metformin and DPP4 combination, OTHERS = all other anti-hyperglycaemic drugs.

Multiple patterns emerge both over time and as treatment stages progress. From 2012 to 2021, in the second treatment stage, the proportion of patients receiving SU decreased from 40% to 3%, DPP4 from 23% to 8%, combination products containing metformin and DPP4 from 15% to 4%, and insulin from 7% to 3%. During the same period, the use of GLP1 increased from 9% to 35%, and SGLT2 from 0% to 43%. GLP1, SGLT2, and insulin became the primary drugs introduced from the third-line treatment stage onwards. The proportion of patients using insulin increased substantially as the stages progressed, but over time, the proportion of patients using SGLT2, and especially GLP1, increased to almost rival that of insulin. There was insufficient data for the fifth- and sixth-line treatments in 2012.

### 4.5 Treatment Pathways

The top 20 pathways of treatment are presented in appendix A1. The most frequent drug treatment pathways contain two or three drugs, and between the two inclusion periods, the proportion of paths containing DPP4 decreased, whereas pathways with SGLT2 and GLP1 increased. In both periods, the most frequent paths of treatment starts with metformin, followed by SGLT2, GLP1, or DPP4. These three paths account for 13.8%, 9.5%, and 7.2% in the early and 28.2%, 24.7%, and 5.8% in the late, out of all paths for all individuals, respectively. While “metformin → SGLT2” remains the most popular path on a class level in both periods, “metformin → injectable semaglutide” is the most frequent path in the late period. The unique path in the late period where 3.8% have the path “GLP1 → GLP1”, is mainly due to people moving from liraglutide to injectable semaglutide.

## 5 Conclusions

### 5.1 Main findings

This study identified several changes in treatment patterns and patient characteristics between 2012 and 2021, for a nationwide cohort of people with type 2 diabetes on who received second-line treatment in the study period. Four key treatment pattern changes were found. *First*, the rapid increase in utilization of GLP1 and SGLT2, which by 2021 approached that of metformin, began prior to the publication of the 2018 guidelines (1,3). An overall increase in utilization of nNIADs has previously been reported in Denmark (5,6) and other Western countries (8–12,23,34). The literature on the subject generally agrees that drug utilization should be higher (8–13,22,23,34). Only one study from the US (21) suggests that the 2018 guidelines are being followed. While some studies may suggest that utilization could be higher, this study indicates that it is high in Denmark. *Second*, the first-line use of SGLT2 and GLP1 was low until a sharp increase occurred in 2021, especially for GLP1. A Danish study (6) found that in 2021, 20% of all first-line glucose-lowering therapies prescribed by general practitioners were GLP1, and 59% of hospital-prescribed first-line glucose-lowering therapies were SGLT2. This study also used data from the DNPD but included all users of any glucose-lowering prescriptions aged 18 years or older. Although several factors may contribute to this increase, adherence to personalized treatment guidelines is likely one of them. In Canada, between 2016 and 2020, first-line use was 10% for GLP1 and 1.6% for SGLT2 (7), while a study from the US (34) indicated low use of these drugs as first-line therapy, but a steady increase since 2017. The first-line use of SGLT2 and GLP1 reported in the present study may be a conservative estimate of this increasingly common international phenomenon. *Third*, investigating changes over time in treatment stages revealed notable changes in treatment patterns. Mainly, SGLT2 and GLP1 were introduced increasingly from second- to sixth-line. Combined, they surpassed insulin at the later treatment stages and as time progressed. If later treatment stages indicate progressed diabetes, as more different drugs have been introduced to the patient, this represents a significant change in treatment practices. SGLT2 and especially GLP1 are being used more frequently over time. The increased use of SGLT2 and GLP1 at later stages could be due to guideline-induced changes in prescribing behavior, suggesting that more options are explored before initiating insulin, or that insulin initiation can be postponed due to better glycemic management from using GLP1 and SGLT2. Extending the time until insulin initiation has long been a goal in diabetes treatment (1,35), indicating that this could be due to an improvement in the treatment of patients with type 2 diabetes. *Fourth*, the three most frequent treatment stage pathways for patients were metformin followed by either SGLT2, GLP1, or DPP4. Between the two periods, the proportion these treatment stage pathways doubled from 30.5% to 58.8%. This could indicate that adherence to the nNIADs was high. An additional observation was the 3.8% in the late period on the “GLP1→GLP1” treatment stage pathway, which may have represented those using GLP1 for weight loss instead of type 2 diabetes treatment.

### 5.2 Implications and Future Perspectives

This study underscores the benefits of incorporating multiple elements into the monitoring of treatment patterns. Denmark’s efforts have been extensive, including national and provider-level monitoring (6), expenditure tracking (5), and a focus on the uptake of nNIADs for their cardiovascular disease (CVD) indications (36). Our approach of concentrating on a specified cohort of individuals with type 2 diabetes provides valuable insights into evolving treatment patterns, which could inform more precise clinical recommendations. Integrating all these approaches would be ideal and is possible due to the extensive and high quality registry data in Denmark. The rapid uptake of the nNIADs, combined with their high prices necessitates these monitoring efforts. Recent health economic literature indicates that the benefits of SGLT2 and GLP1 outweigh their costs for second-line treatment (37). One study concludes that as first-line treatment, price reductions are necessary to achieve cost-effectiveness (38), and the UK have deemed GLP1 to be not cost-effective at their current price levels (39). This can change in the near future, as a generic version for liraglutide is expected in 2024 (40), and patents for sitaglipin and empagliflozin expires soon.

### 5.3 Strengths and Limitations

Our study benefits from the use of nationwide prescription data, which offers a reliable tracking of real-world treatment patterns and minimizes selection and information bias. However, several important limitations must be considered. First, we did not differentiate between GLP1 products used specifically for weight loss and those for type 2 diabetes, which could have led to some inclusion of individuals without a T2D diagnosis, although the amount is likely to be small, and possibly encapsulated by the 1370 or more individuals going from liraglutide to semaglutide, see appendix A1. Due to the inclusion criteria for the cohort, and the shifting trends in first-line prescribing practice, we decided the inclusion criteria were sufficient in selecting the target cohort. Second, the identification of treatment stages could have been more precise by establishing clear criteria for treatment intensification and systematically tracking medications at each stage, which would have provided a more accurate understanding of treatment progression. Third, our reliance solely on prescription data, without including hospital admission diagnoses or paraclinical data, may have missed additional context that could improve the precision of identifying patients with type 2 diabetes. However, the completeness of prescription data provides a high level of reliability at the nationwide scale.

### 5.4 Concluding remarks

The drug treatment patterns for a cohort of individuals with type 2 diabetes on second-line treatment in Denmark have changed substantially. SGLT2 and GLP1 are surpassing all other drug classes in utilization and may have led to decreased insulin use at later stages of treatment for this cohort. Their use as first-line options are evolving rapidly. This rapid change in treatment practices for type 2 diabetes carries significant benefits but necessitates intensified monitoring efforts to ensure that prescribing practices are consistent with treatment guidelines.

## Supporting information

RECORD statement

## Data Availability

Underlying micro-data are not available, as they are protected on a research server. All relevant aggregate data is made available in the manuscript or appendices.

## 5.5 Acknowledgements

## 5.5.1 Funding and assistance

This study was supported by Steno Diabetes Center North Denmark.

## 5.5.3 Author contributions and Guarantor Statement

HVBL was responsible for the conception of the study question, analysis, presenting, and describing data, and writing the manuscript drafts. SPJ, PV, and FWU provided ideas for the conception of the study question, generated ideas regarding focus areas of the analysis, interpreted the results and provided feedback to the manuscript. MHJ has interpreted the results and provided feedback to the manuscript. All authors agree to be responsible for all aspects of the work and have read and approved the final manuscript.

## 5.4.3 Prior presentation

This study has not been presented previously.

