## Supplemental Files for "Trends in anti-hyperglycaemic drug usage among Danish type 2 diabetes patients on second-line treatment and beyond: A nationwide cohort study"

### Appendix A1

*Top 20 treatment stage pathways for the cohort, with the percentage of individuals with each path out of the total amount in each time period. n = total amount of unique paths, N = total amount of individuals. MET = metformin, SU = Sulfonylurea, DPP4 = dipeptidyl peptidase-4 inhibitors, GLP1 = glucagon-like peptide-1 receptor agonists, SGLT2 = sodium glucose co-transporter-2 inhibitor, TZD = thiazolidinedions, AGI = alpha-glucosidase inhibitor, INS = insulin, METandDPP4 = metformin and DPP4 combination, semaglutide-i = injectable semaglutide, semaglutide-o = oral semaglutide.*

| Drug class | | | | Drug product | | | |
|---|---|---|---|---|---|---|---|
| Early (n=4,321,N =51,796) | | Late (n=552,N =14,392) | | Early (n=9,861,N=51,7 96) | | Late (n=1,211,14,392) | |
| PATH | Count (%) | PATH | Count (%) | PATH | Count (%) | PATH | Count (%) |
| MET→SGLT2 | 7,170 (13.84) | MET→SGLT2 | 4,057 (28.19) | MET→empagliflozin | 4,652 (8.98) | MET→semaglutide-i | 2,571 (17.86) |
| MET→GLP1 | 4,904 (9.47) | MET→GLP1 | 3,560 (24.74) | MET→semaglutide-i | 3,041 (5.87) | MET→empagliflozin | 2,411 (16.75) |
| MET→DPP4 | 3,724 (7.19) | MET→DPP4 | 841 (5.84) | MET→dapagliflozin | 2,379 (4.59) | MET→dapagliflozin | 1,564 (10.87) |
| MET→METandDPP4 | 2,329 (4.5) | MET→SGLT2→GLP1 | 714 (4.96) | MET→sitagliptine | 2,321 (4.48) | MET→semaglutide-o | 774 (5.38) |
| MET→SGLT2→GLP1 | 2,152 (4.15) | GLP1→GLP1 | 546 (3.79) | MET→glimepiride | 1,770 (3.42) | MET→sitagliptine | 579 (4.02) |
| MET→SU | 2,107 (4.07) | MET→SU | 376 (2.61) | MET→metANDsitagliptine | 1,370 (2.64) | liraglutide→semaglutide-i | 425 (2.95) |
| MET→DPP4→SGLT2 | 1,283 (2.48 | MET→METandDPP4 | 359 (2.49) | MET→liraglutide | 1,022 (1.9 | MET→empagliflozin→semaglutide-i | 324 (2.25) |

| Drug class | | | | Drug product | | | |
|---|---|---|---|---|---|---|---|
| | ) | | | | 7) | | |
| MET→METandDPP4→SGLT2 | 1,202 (2.32) | MET→METandSGLT2 | 292 (2.03) | MET→empagliflozin→semaglutide-i | 946 (1.83) | MET→glimepiride | 313 (2.17) |
| MET→DPP4→GLP1 | 789 (1.52) | MET→GLP1→SGLT2 | 217 (1.51) | MET→metANDvildagliptine | 894 (1.73) | MET→metANDsitagliptine | 237 (1.65) |
| MET→GLP1→GLP1 | 735 (1.42) | MET→DPP4→SGLT2 | 169 (1.17) | MET→linagliptine | 864 (1.67) | MET→liraglutide | 205 (1.42) |
| MET→METandSGLT2 | 735 (1.42) | MET→SGLT2→DPP4 | 156 (1.08) | MET→semaglutide-o | 802 (1.55) | MET→metANDempagliflozin | 180 (1.25) |
| MET→SGLT2→DPP4 | 586 (1.13) | GLP1→MET | 142 (0.99) | MET→liraglutide→semaglutide-i | 658 (1.27) | MET→linagliptine | 175 (1.22) |
| MET→SU→SGLT2 | 585 (1.13) | SGLT2→MET | 135 (0.94) | MET→sitagliptine→empagliflozin | 573 (1.11) | MET→dapagliflozin→semaglutide-i | 157 (1.09) |
| MET→METandDPP4→GLP1 | 570 (1.1) | MET→SGLT2→METandSGLT2 | 122 (0.85) | MET→dapagliflozin→semaglutide-i | 496 (0.96) | MET→empagliflozin→semaglutide-o | 126 (0.88) |
| MET→GLP1→SGLT2 | 538 (1.04) | MET→DPP4→GLP1 | 113 (0.79) | MET→metANDsitagliptine→empagliflozin | 457 (0.88) | MET→metANDvildagliptine | 111 (0.77) |
| MET→DPP4→SGLT2→GLP1 | 525 (1.01) | MET→INS→GLP1 | 103 (0.72) | MET→metANDempagliflozin | 451 (0.87) | MET→metANDdapagliflozin | 108 (0.75) |
| MET→METandDPP4→SGLT2→GLP1 | 475 (0.92) | MET→INS→SGLT2 | 83 (0.58) | MET→sitagliptine→semaglutide-i | 389 (0.75) | dapagliflozin→MET | 82 (0.57) |
| MET→DPP4→METandDPP4 | 380 (0.73) | MET→METandDPP4→SGLT2 | 81 (0.56) | MET→vildagliptine | 361 (0.7) | MET→canagliflozin | 80 (0.56) |
| MET→SU→GLP1 | 350 (0.68) | MET→GLP1→GLP1 | 73 (0.51) | MET→sitagliptine→dapagliflozin | 335 (0.65) | MET→semaglutide-i→empagliflozin | 77 (0.54) |
| MET→SU | 333 | MET→S | 67 | MET→glimepiri | 329 | MET→empaglifl | 75 |

| Drug class | | | | Drug product | | | |
|---|---|---|---|---|---|---|---|
| →DPP4 | (0.64<br>) | GLT2→I<br>NS | (0.47<br>) | de→empagliflozi<br>n | (0.6<br>4) | ozin→metANDe<br>mpagliflozin | (0.52<br>) |
